# Digital droplet PCR accurately quantifies SARS-CoV-2 viral load from crude lysate without nucleic acid purification

**DOI:** 10.1101/2020.09.02.20186023

**Authors:** Harish N. Vasudevan, Peng Xu, Venice Servellita, Steve Miller, Leqian Liu, Allan Gopez, Charles Y. Chiu, Adam R. Abate

## Abstract

The COVID-19 pandemic caused by the SARS-CoV-2 virus motivates diverse diagnostic approaches due to the novel causative pathogen, incompletely understood clinical sequelae, and limited availability of testing resources. Given the variability in viral load across and within patients, absolute viral load quantification directly from crude lysate is important for diagnosis and surveillance. Here, we investigate the use of digital droplet PCR (ddPCR) for SARS-CoV-2 viral load measurement directly from crude lysate without nucleic acid purification. We demonstrate ddPCR accurately quantifies SARS-CoV-2 standards from purified RNA and multiple sample matrices, including commonly utilized universal transport medium (UTM). In addition, we find ddPCR functions robustly at low input viral copy numbers on nasopharyngeal swab specimens stored in UTM without upfront RNA extraction. We also show ddPCR, but not qPCR, from crude lysate shows high concordance with viral load measurements from purified RNA. Our data suggest ddPCR offers advantages to qPCR for SARS-CoV-2 detection with higher sensitivity and robustness when using crude lysate rather than purified RNA as input. More broadly, digital droplet assays provide a potential method for nucleic acid measurement and infectious disease diagnosis with limited sample processing, underscoring the utility of such techniques in laboratory medicine.

## Introduction

The COVID-19 pandemic represents a global health emergency with over 6 million cases of SARS-CoV-2 infection and 359,000 deaths reported to date.^1^ Accurate diagnostic testing is essential, yet false negative results persist with current COVID-19 tests, particularly during early stages of infection prior to symptom onset.^2^ In addition, SARS-CoV-2 viral load varies throughout an individual patient’s disease course.^3^ Indeed, repeat testing of patient specimens can yield disparate viral loads without a change in clinical status, increasing the false negative rate and complicating patient management. Given the key role of diagnostic testing in guiding public health initiatives and strong relationship between viral load and transmissibility,^4^ these observations underscore the importance of accurate approaches for SARS-CoV-2 quantification.

The gold standard for COVID-19 diagnosis is quantitative reverse transcriptase PCR (qRT-PCR) as recommended by the US Centers for Disease Control and Prevention (CDC).^5^ Although robust in many settings,^6^ qRT-PCR is limited by its reliance on a standard curve, sensitivity to inhibitors in clinical samples, and inconsistent performance at low concentrations. Furthermore, current COVID-19 qRT-PCR tests require multiple upstream processing steps, including sample collection, viral lysis, and RNA purification, a workflow limited by shortages in laboratory supplies and RNA extraction kits leading to bottle necks in COVID-19 testing. To mitigate these shortcomings, numerous tests are under development, including loop-mediated isothermal amplification (LAMP)^7,8^ and CRISPR-based assays.^9,10^ Although promising, these methods generally require upfront nucleic acid purification. Experimental approaches attempt to analyze crude lysate directly,^11,12^ but the resulting viral load measurements are not quantitative and overall assay performance remains unknown. Thus, a robust method to quantify SARS-CoV-2 viral load directly from crude lysate would both simplify testing and provide additional information to potentially guide clinical management.

Digital droplet PCR quantifies target nucleic acid sequences using many partitioned reactions. In contrast to qRT-PCR, in which concentrations are inferred from amplification rates relative to a standard curve, ddPCR cycles the sample to endpoint, after which target molecules are counted directly by enumerating positive droplets. This approach provides several advantages over qRT-PCR, including more precise measurements and absolute quantification without the need for a standard curve.^13,14^ Moreover, ddPCR can detect a variety of viral pathogens including human immunodeficiency virus (HIV),^15^ cytomegalovirus (CMV),^16^ and human herpes virus 6 (HHV-6).^17^ In COVID-19 patients, ddPCR of purified RNA extracts demonstrates advantages for diagnosis and monitoring, particularly in patients exhibiting low viral load.^18–20^ Although recent studies suggest ddPCR may be resistant to lysate-based inhibition,^21^ fidelity from crude viral lysate without RNA extraction for SARS-CoV-2 viral load quantification remains unknown.

Here, we show that digital droplet PCR (ddPCR) enables accurate SARS-CoV-2 RNA quantification from viral lysate without nucleic acid purification. It provides absolute viral counts from crude lysate, obviating the need for a standard curve while being resistant to reaction inhibition. In addition, we find that ddPCR, but not qRT-PCR, yields accurate measurement of SARS-CoV-2 viral load when applied directly to crude lysate without RNA extraction. In addition, ddPCR from crude lysate provides comparable viral load estimates to qRT-PCR from purified nucleic acid. Finally, ddPCR detects SARS-CoV-2 RNA in a clinically suspicious case negative by conventional qRT-PCR. Taken together, these data indicate that ddPCR robustly quantifies SARS-CoV-2 RNA from crude viral lysate, thus representing a complementary approach to conventional qRT-PCR in clinically ambiguous scenarios where ultra-high sensitivity is needed or when RNA purification cannot be incorporated within the diagnostic workflow.

## Materials and Methods

### SARS-CoV-2 quantitative reverse transcriptase polymerase chain reaction

N1 and N2 nucleocapsid primers from the CDC assay were obtained from Integrated DNA Technologies (IDT). PCR cycling conditions including reverse transcription were run per the CDC EUA-approved protocol5 with the Promega GoTaq Probe 1-Step RT-qPCR system (Catalog # A6120). In brief, a 20 μL reaction comprising 10 μL GoTaq, 0.4 μL GoScript, 1.5 μL primer master mix, 3.1 μL water, and 5 μL input RNA (or crude lysate) was reverse transcribed by incubating at 45° C for 15 minutes followed by 95° C for 2 minutes per CDC protocol. The reaction was then thermocycled for 45 cycles of 95° C for 20 seconds (denaturation) and 55° C for 30 seconds (annealing) on either the ABI 7500 Fast DX (Applied Biosystems) or the QuantStudio 5 Real-Time PCR System (Thermo Fisher).

### Digital droplet polymerase chain reaction

The N1 nucleocapsid primers from the CDC assay were used for all ddPCR data with the 1-step RT-ddPCR Advanced Kit for Probes (Bio-Rad Catalog # 1864022). In brief, a 20 μL reaction comprising 2 μL reverse transcriptase, 1 μL DTT, 1.5 μL primer master mix, 5.5 μL water, and 5 μL input RNA (or crude lysate) was used for each sample. Droplets were generated with the QX200 Droplet Digital PCR System (Bio-Rad), sealed, and reverse transcription was performed by incubating at 50° C for 60 minutes followed by 95° C for 10 minutes per manufacturer’s protocol. The reaction was then thermocycled with the same conditions as used for the CDC EUA-approved bulk qRT-PCR assay described above, and then read with the QX200 Droplet Reader (Bio-Rad) with thresholds between positive and negative drops set by QuantaSoft Software and confirmed by manual inspection. For qualitative assessment of fluorescence, thermocycled drops were imaged using the EVOS Cell Imaging System (Thermo Fisher).

### SARS-CoV-2 RNA and viral standard preparation

In vitro transcribed SARS-CoV-2 RNA standards obtained from Twist biosciences (Catalog #102916) were used for standard curve calculation across qRT-PCR and ddPCR. Replication defective virus was obtained from Seracare via the AccuPlex SARS-CoV-2 Reference Material Kit (Catalog #0505-0126), and purified RNA was generated from these standards with the Qiagen DSP Viral RNA Mini Kit (Catalog #61904) as described in the CDC EUA approved protocol. For crude lysate, samples were processed either using the QuickExtract lysis buffer (Lucigen Catalog # QE09050) as previously described,11 simply heated for 5 minutes at 95° C, or added directly to UTM for downstream analysis.

### Human nasopharyngeal swab sample collection and preparation from COVID positive patients

Clinical nasopharyngeal swab samples from patients infected with SARS-CoV-2 were collected in UTM and acquired by the Chiu laboratory with approval of the University of California San Francisco (UCSF) Institutional Review Board (IRB). The approved study was a no-subject contact biobanking protocol using remnant clinical samples with waiver of consent under approval from the University of California San Francisco (UCSF) Institutional Review Board (IRB). All experiments were performed in accordance with relevant guidelines and regulations per approved UCSF IRB protocol. RNA purification and crude lysis extraction was performed as described above. For qRT-PCR from purified RNA, samples were diluted 1:4 given they were concentrated two-fold during purification and not mixed 1:1 with Quick Extract buffer in order to provide direct comparison to crude lysis conditions. Of the 33 original clinical samples obtained for analysis, 32 had sufficient material for qRT-PCR from both purified RNA and crude lysate while 22 had sufficient material for adequate dropmaking and subsequent comparison across all three assays (qRT-PCR from purified RNA, qRT-PCR from crude lysate, and ddPCR from crude

## Results

We sought to compare the performance of conventional qRT-PCR and ddPCR for SARS-CoV-2 detection (Figure 1). The standard qRT-PCR workflow for SARS-CoV-2 testing requires sample collection via nasopharyngeal swab, placement of swabs into sterile tubes containing universal transport medium (UTM), and nucleic acid extraction to obtain purified RNA free of PCR inhibitors (Figure 1, top workflow). The subsequent qRT-PCR result is thus critically dependent on the amount and quality of template RNA extracted, potentially resulting in false negatives at low input viral copy numbers. A simpler approach would be to omit the RNA extraction step, thus performing quantification directly on cell lysate. However, inhibitors present in unpurified cell lysate decrease reaction efficiency and interfere with accurate quantification, potentially leading to false negatives. We hypothesize ddPCR circumvents this shortcoming by partitioning the target nucleic acid and sequestering inhibitory molecules in separate droplets, permitting normal reaction kinetics and accurate viral quantification (Figure 1, bottom workflow). From a practical perspective, such an approach would simplify the overall workflow and obviate the need for a standard curve.

**Figure 1.**
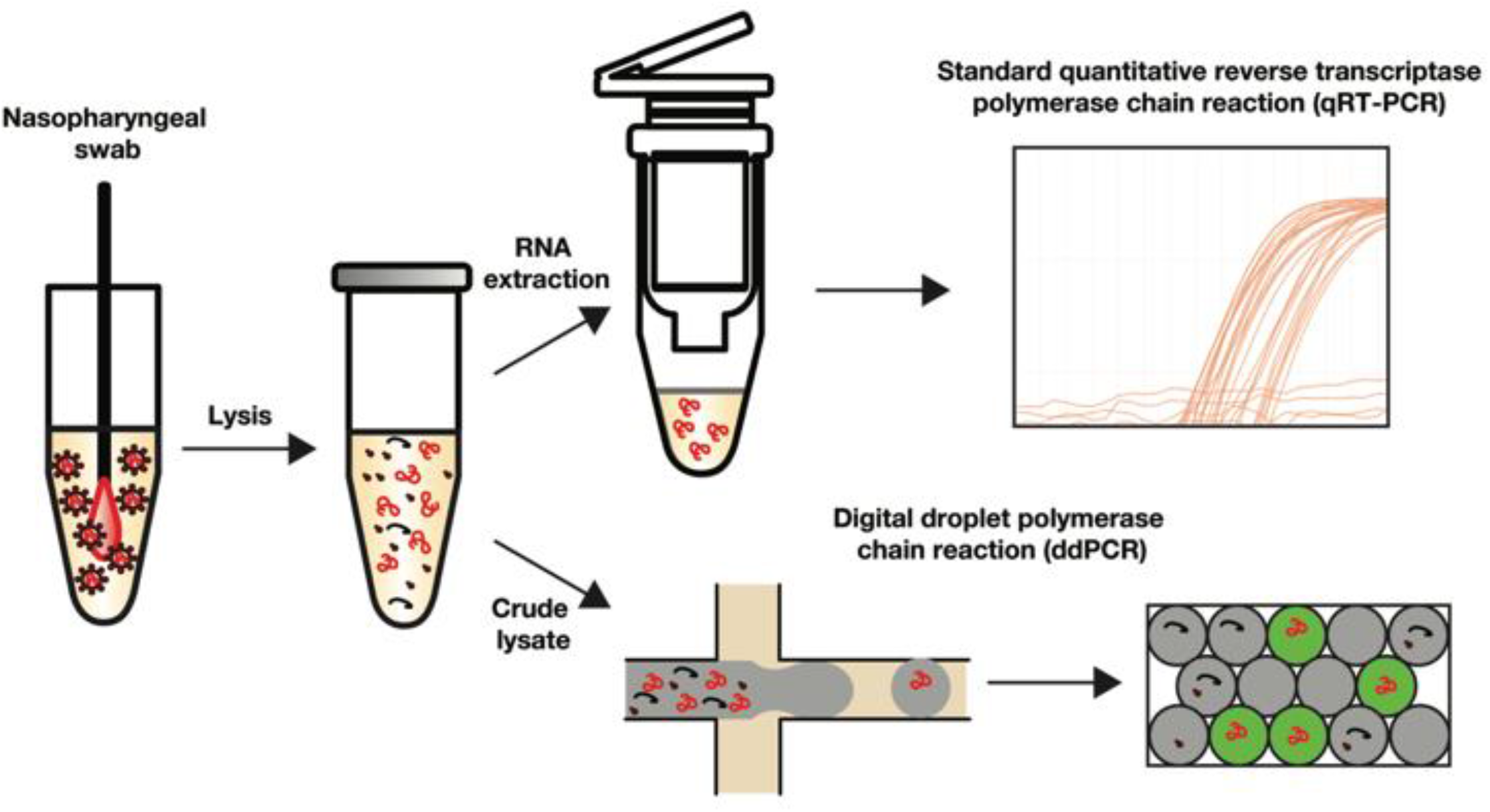
Comparison of the CDC SARS-CoV-2 detection assay requiring RNA extraction followed by bulk qRT-PCR (top workflow) versus our ddPCR approach directly on crude lysate (bottom workflow) Red molecules represent target nucleic acids while black molecules represent viral proteins and other reaction inhibitors present in unpurified cell lysate. Digital droplet PCR potentially improves viral load quantification by sequestering reaction inhibitors in separate droplet from target nucleic acid sequences.

We first adapt the nucleocapsid N1 primer used in the CDC qRT-PCR assay for ddPCR and evaluate our ddPCR assay on purified, in vitro transcribed SARS-CoV-2 RNA standards (Twist Biosciences, Catalog #102916). Visualization of droplet fluorescence reveals a clear qualitative difference between positive and negative drops (Figure 2a) with minimal background fluorescence with a no template control (Figure 2a’). Quantitative analysis of fluorescence intensity similarly shows clear separation with an approximately two-fold increase in fluorescence of positive compared to negative drops (Figure 2b), again with minimal false positives in the no-template control (Figure 2b’). To confirm the linearity and dynamical range of ddPCR, we compare SARS-CoV-2 RNA standards analyzed by conventional qRT-PCR (Figure 2c) versus ddPCR (Figure 2d). Both bulk qRT-PCR (r_2_ = 0.96) and ddPCR (r_2_ = 0.92) robustly quantify input RNA across the tested range and exhibit similar limits of detection at 10 copies per reaction, consistent with published reports.^18–20^

**Figure 2.**
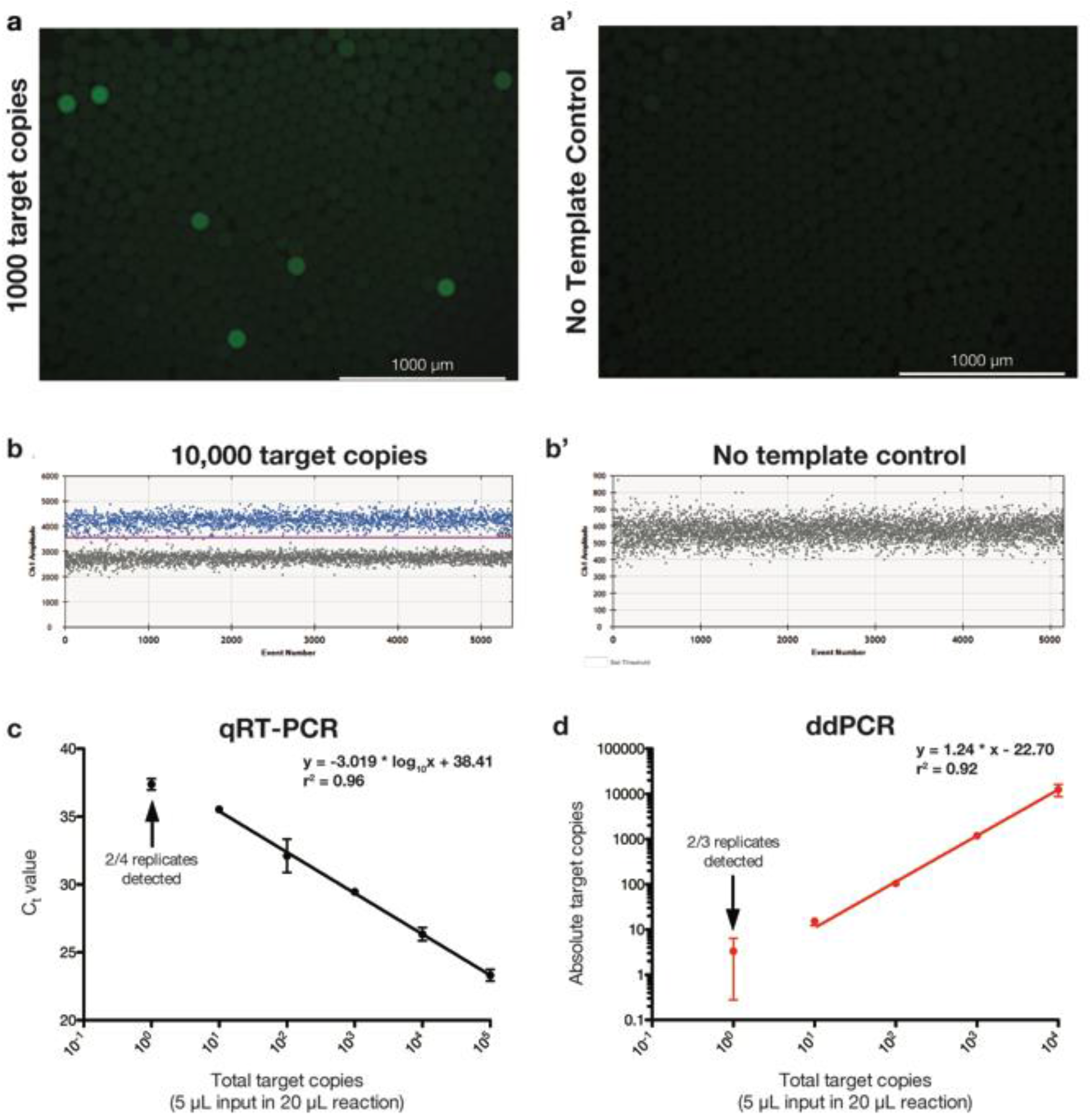
Digital droplet PCR (ddPCR) reliably quantifies SARS-CoV-2 RNA standards. (a) Fluorescence microscopy images of ddPCR using CDC N1 primer set with 1000 input copies of SARS-CoV-2 RNA and (a’) no template control. (b) Fluorescence intensities as measured by BioRad QX200 Droplet Reader for 10,000 input target copies and (b’) no template control. (c) Quantitative reverse transcription PCR (qRT-PCR) standard curve using CDC N1 primer set and SARS-CoV-2 RNA shows expected linear relationship between input viral copy number and cycle threshold (Ct) value with an estimated limit of detection (LoD) of 10 copies per reaction. (d) ddPCR standard curve for SARS-CoV-2 RNA reveals accurate viral copy number measurement with an estimated LoD of 10 copies per reaction.

Unlike purified standards, crude lysate from patient samples contains target viral nucleic acids mixed with numerous inhibitory molecules. Thus, novel approaches aiming to directly analyze patient samples without nucleic acid purification implement lysis strategies, such as heating input samples or utilizing extraction buffers that stabilize reaction enzymes and neutralize inhibitors. To determine how such workflows impact assay performance, we compare qRT-PCR and ddPCR quantification of SeraCare SARS-CoV-2 viral standards using three crude lysis workflows without RNA purification: (1) incubation at 95 °C for 5 minutes directly (heat lysis), (2) addition of QuickExtract buffer in a 1:1 ratio followed by incubation at 95°C for 5 minutes,11 or (3) directly from UTM. Conventional qRT-PCR detects SeraCare SARS-CoV-2 standards across all three crude lysis conditions as well as purified RNA at both 125 and 12.5 input copies per reaction using an assay cycle threshold (Ct) of less than 40 per CDC recommendations (Figure 3a). However, we observe increased Ct values for all tested crude lysate conditions in the absence of upfront RNA purification except for QuickExtract buffer at 125 input copies, suggesting decreased efficiency when amplifying template from crude lysate by qRT-PCR. In contrast, ddPCR accurately and reproducibly quantifies input viral copy number across all conditions (Figure 3b). Intriguingly, UTM inhibits bulk qRT-PCR but does not affect ddPCR, suggesting ddPCR may be useful for retrospective analysis of patient samples stored in UTM. No false positives are present in the QuickExtract buffer samples analyzed by ddPCR, suggesting adequate digestion of background nucleic acid. When compared to estimated SARS-CoV-2 viral load by relative quantification from qRT-PCR standard curves, ddPCR offers more accurate viral load measurement both at 125 input copies per reaction (Figure 3c) and 12.5 input copies per reaction (Figure 3d). Taken together, this shows that ddPCR, but not qRT-PCR, accurately quantifies SARS-CoV-2 viral load from crude lysate without RNA purification at low input copy numbers and in the presence of UTM.

**Figure 3.**
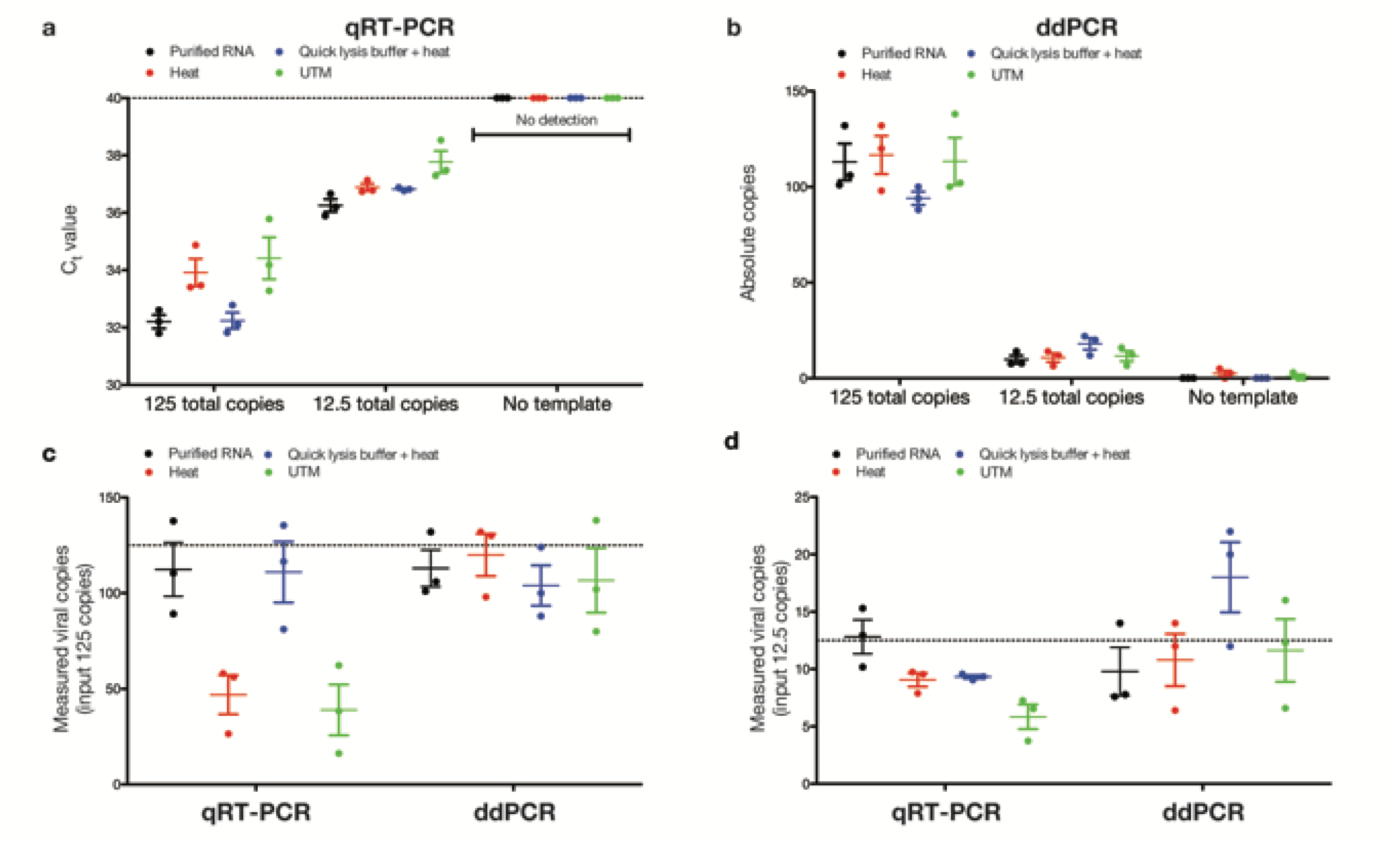
Digital droplet PCR (ddPCR) provides robust SARS-CoV-2 viral load measurement across crude lysis conditions. (a) Quantitative reverse transcription PCR (qRT-PCR) of Seracare SARS-CoV-2 viral standards across lysis conditions reveals decreased PCR efficiency at low copy numbers and in presence of universal transport medium (UTM). (b) ddPCR of Seracare SARS-CoV-2 viral standards across lysis conditions reveals accurate viral load quantification across all tested conditions. (c) Comparison of relative quantification by qRT-PCR and absolute quantification by ddPCR at 125 input copies and (d) 12.5 input copies shows decreased viral load measurements by qRT-PCR in the presence of UTM.

To evaluate both qRT-PCR and ddPCR methods for SARS-CoV-2 RNA quantification from patient samples collected by nasopharyngeal swab and stored in UTM, we test 33 clinical specimens collected from patients suspected of having COVID-19. When comparing qRT-PCR from purified RNA versus crude lysate, the Ct values obtained following amplification from crude lysis processed with QuickExtract buffer are consistently increased compared to Ct values from purified RNA, suggesting decreased reaction efficiency and subsequently, inaccurate estimate of viral load (Figure 4a). In contrast, absolute quantification of SARS-CoV-2 viral load by ddPCR from crude lysate shows consistency with relative quantification by qRT-PCR from purified RNA rather than viral load estimates obtained by qRT-PCR from crude lysate (Supplemental Figure 1a). Direct inspection of fluorescence intensities confirms clear separation between positive and negative drops in patient samples without false positives in the no-template control (Supplemental Figure 1b), consistent with robust detection by ddPCR. When estimating bias by calculating the difference in viral load quantification relative to qRT-PCR from purified RNA, qRT-PCR from crude lysate shows increased differences compared to ddPCR from crude lysate at both low input viral load < 10^3^ copies per reaction (Figure 4b) but not high input viral load > 10^3^ copies per reaction (Figure 4b’). Finally, relative quantification by qRT-PCR directly from crude lysate, while positively correlated with measurements from purified RNA (y = 0.69x – 190, r_2_=0.53), consistently underestimates SARS-CoV-2 viral load in clinical patient samples, particularly at lower input viral load where qRT-PCR variance is increased (Figure 4c). In contrast, absolute quantification by ddPCR demonstrates strong correlation with estimates from qRT-PCR performed from purified RNA (y = 0.80x - 172, r_2_=0.92), highlighting the improved assay fidelity with ddPCR compared to qRT-PCR when working from crude lysate (Figure 4d). In sum, ddPCR demonstrates better correlation with the current gold standard of qRT-PCR from purified RNA for quantitation of SARS-CoV-2 viral load.

**Figure 4.**
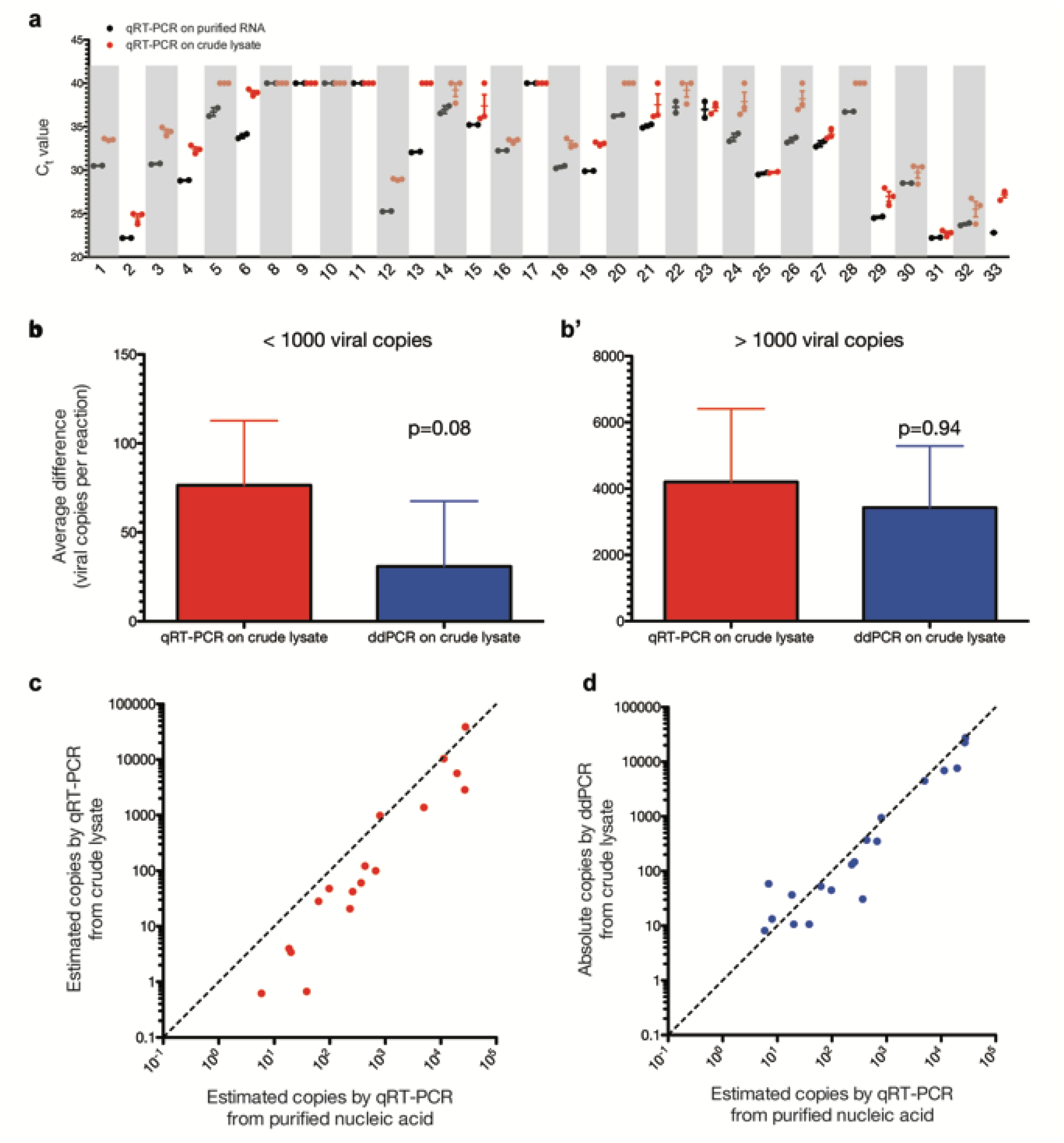
Digital droplet PCR (ddPCR), but not bulk qRT-PCR, accurately quantifies SARS-CoV-2 viral load from patient nasopharyngeal samples without nucleic acid isolation. (a) Ct values for qRT-PCR from purified RNA (black) versus RNA prepared from crude lysate (red) shows decreased PCR efficiency without upfront nucleic acid purification of patient samples as evidenced by increase Ct values (n=32 samples). (b) Average difference in viral load between qRT-PCR from purified RNA versus either qRT-PCR from crude lysate (red) or ddPCR from crude lysate (blue) demonstrates increased bias with qRT-PCR compared to ddPCR for low viral load (< 1000 copies / reaction, p=0.08) but not (b’) high viral load (> 1000 copies per reaction, p=0.94). (c) Relative quantification of viral load by qRT-PCR from purified RNA versus crude lysate demonstrates systematic underestimation of viral load. (d) Absolute quantification of viral load by ddPCR from crude lysate shows strong correlation with relative quantification by qRT-PCR from purified RNA. In panels (c) and (d), graphed y=x line provides a reference for perfect agreement between the two assays. All crude lysis was carried out via 1:1 dilution QuickExtract buffer followed by heating at 95° C for 5 minutes as described in text.

## Discussion

The COVID-19 pandemic underscores the importance of diagnostic testing for novel pathogens with assays that minimize sample processing and reagent requirements while maintaining accuracy. Here, we show ddPCR allows accurate SARS-CoV-2 quantification from crude lysate while conventional bulk qRT-PCR, the current gold standard, appears more sensitive to inhibition. When analyzing crude lysate from patient samples, the result is higher rates of false negatives by qRT-PCR with viral loads near the limit of detection. In contrast, ddPCR is robust to inhibition and accurately quantifies viral load without nucleic acid purification. While the clinical relevance of patients harboring virus at low titers with C^t^ values > 36 remains unclear, robust detection of such individuals may facilitate identification of asymptomatic spreaders, monitoring disease progression, and evaluating the efficacy of antiviral therapy.

While our results are consistent with previous reports that ddPCR is more resistant to reaction inhibition than bulk qPCR,^21^ the underlying mechanism remains poorly understood. One hypothesis is that sequestration of inhibitors in empty droplets facilitates more efficient PCR amplification in droplets containing template. In this scenario, the fidelity of droplet assays in crude lysate would likely be independent of the specific molecular biology used for nucleic acid detection, and thus, such resistance to inhibition may extend to other assays such as isothermal and CRISPR-based target detection. Indeed, droplet LAMP for influenza exhibits increased resistance to reaction inhibition.^21^ Future work investigating the mechanism underlying the improved performance of droplet over bulk assays for other SARS-CoV-2 detection methods from crude lysate will be important to extend the generalizability of our findings.

Detection of low titer SARS-CoV-2 RNA by ddPCR in patients negative by the ‘gold standard’ clinical qRT-PCR assay raises a potential concern of false positive results. While current qRT-PCR COVID-19 testing appears prone to false negatives,^22–25^ it nevertheless remains possible ddPCR demonstrates the opposite predilection for false positives. We attempt to control for this by including bona fide no template controls for all assay runs. Furthermore, our finding of similar limit of detection but improved precision with ddPCR compared to qRT-PCR is consistent with prior work in cancer.^26^ Importantly, comparison of ddPCR to qRT-PCR from purified nucleic acid extracts in larger COVID-19 patient cohorts similarly demonstrates detection of SARS-CoV-2 RNA by ddPCR in patients negative by qRT-PCR.^19,20^ Nevertheless, additional prospective comparisons of qRT-PCR and ddPCR in larger patient cohorts are needed to confirm these observations.

Our work highlights the potential advantages of ddPCR for SARS-CoV-2 diagnosis, particularly to obtain quantitative results from crude lysate, thus obviating the need for nucleic acid purification. Our results motivate further validation of droplet assays for COVID-19 in the clinical laboratory setting for patients at high clinical suspicion of COVID-19 infection despite negative qRT-PCR test results. While the clinical relevance of patients harboring virus at low titers with Ct values > 36 remains unclear, robust detection of such individuals may facilitate identification of asymptomatic spreaders, monitoring disease progression, and evaluating the efficacy of antiviral therapy. Furthermore, monitoring low viral loads may potentially be useful in the public health arena for disease monitoring in presymptomatic or asymptomatic cases with low viral loads^27,28^ or across point-of-care applications where minimal sample processing is essential. More broadly, the present data support the role of ddPCR as a potential alternative to bulk qPCR in settings requiring nucleic acid quantification at low input copy numbers without upfront nucleic acid extraction.

## Data Availability

All data is available as primary or supplemental figures in the manuscript.

## Data availability

Not applicable, all presented data in manuscript.

## Acknowledgments

We thank the patients and their families at UCSF without whom collecting and providing this aggregate data would not have been possible.

## Contributions

HNV, PX, CYC and ARA designed the study. HNV, PX, LL, and VS performed experiments. HNV, PX, and VS performed data analysis. AG and SM provided samples and technical support. CYC and ARA supervised the study. HNV and ARA prepared the manuscript with input from all authors.

